# The Global Pediatric Diarrhea Surveillance network: Rationale and methods

**DOI:** 10.64898/2026.05.21.26352576

**Authors:** Heidi M. Soeters, Sébastien Antoni, Shilpa S. Iyer, Goitom Weldegebriel, Joseph Biey, Jason M. Mwenda, Gloria Rey-Benito, Claudia Ortiz, Roberta Pastore, Dovile Videbaek, Simarjit Singh, Emmanuel Njambe, Lucky Sangal, Deepak Dhongde, Varja Grabovac, Josephine Logronio, Kamal Fahmy, Amany Ghoniem, George Armah, Francis E. Dennis, Mapaseka L. Seheri, Nonkululeko Magagula, Kebareng Rakau-Nondela, Tulio M. Fumian, Irene T.A. Maciel, Elena Samoilovich, Galina Semeiko, Tintu Varghese, Sarah Thomas, Julie Bines, Dandi Li, Furqan Kabir, Jie Liu, Eric R. Houpt, Rashi Gautam, Sara A. Mirza, Jan Vinjé, Mick N. Mulders, Jacqueline E. Tate, Umesh D. Parashar, James A. Platts-Mills, the Global Pediatric Diarrhea Surveillance network

## Abstract

**Background:** Diarrhea remains a leading cause of child morbidity and mortality worldwide. Improved and ongoing estimates of the etiologies of severe diarrhea, particularly in low- and middle-income countries (LMICs), are crucial to inform the use of current vaccines and other interventions and to help prioritize the development of new vaccines. Producing rigorous longitudinal data on the global burden and etiology of pediatric diarrhea requires a geographically broad surveillance network with standardized epidemiologic, laboratory, and analytic protocols.

**Methods:** We describe the rationale and methods of the Global Pediatric Diarrhea Surveillance (GPDS) network, a World Health Organization (WHO)-coordinated public health surveillance network investigating the etiology of hospitalized diarrhea among children aged <5 years in LMICs. The GPDS network enrolls children hospitalized with diarrhea at 38 sentinel surveillance sites in 31 LMICs across all 6 WHO Regions. Randomly selected stool specimens were tested by TaqMan Array Card quantitative polymerase chain reaction for 16 enteric pathogens previously associated with pediatric diarrhea. GPDS produces estimates of pathogen-specific attributable fractions and incidence of diarrheal hospitalizations at the global, regional, and country levels.

**Conclusions:** As a WHO-coordinated global surveillance network, GPDS evaluates pathogens associated with hospitalized pediatric diarrhea. The network monitor**s** the changing burden of pathogens over time, monitors circulating strains, and generates data to inform decision-making around public health interventions. GPDS also improves global, regional, and country diarrheal disease burden estimates, informs new enteric vaccine development, and potentially provides a platform for future enteric vaccine evaluation.

## Introduction

Despite substantial global progress in the prevention and treatment of enteric diseases, diarrhea remains a leading cause of death and disease in children aged <5 years (1,2). Improved and ongoing estimates of the infectious causes of severe pediatric diarrhea, particularly in low- and middle-income countries (LMICs), are crucial to inform the use of current vaccines and other interventions and to help prioritize the development of new vaccines. Producing rigorous longitudinal data on the global burden and etiology of pediatric diarrhea requires a geographically broad surveillance network with standardized epidemiologic, laboratory, and analytic protocols. Especially in the context of increasing rotavirus vaccine use and impact (3,4), ongoing systematic surveillance is critical.

In this article, we describe the rationale and methods for the Global Pediatric Diarrhea Surveillance (GPDS) network, a World Health Organization (WHO)-coordinated public health surveillance network investigating the etiology of hospitalized diarrhea among children aged <5 years in LMICs starting in 2017.

## Rationale

The WHO has coordinated the Global Rotavirus Surveillance Network (GRSN) since 2008, providing a global source of rotavirus surveillance data among hospitalized children aged <5 years to monitor rotavirus disease burden and circulating genotypes and evaluate rotavirus vaccine effectiveness and impact (5). However, systematically collected and analyzed surveillance data on other infectious causes of diarrhea in hospitalized children has been limited, particularly in LMICs where the burden is highest (1). Advances in quantitative PCR testing have provided enhanced sensitivity and resolution to identify a broad range of pathogens associated with acute diarrhea.

In 2017, WHO leveraged the GRSN sentinel-site surveillance and Regional Reference Laboratory (RRL) system to establish GPDS (6). GPDS investigates the etiology of diarrhea among hospitalized children aged <5 years by using a standardized protocol for quantitative reverse-transcription PCR (qPCR) testing of a broad panel of diarrheal pathogens using the TaqMan Array Card (TAC) platform.

## Objectives

GPDS aims to evaluate pathogens in children hospitalized with diarrhea, which includes monitoring the changing burden of pathogens over time, monitoring circulating strains, and generating country-level data to inform decision-making around public health interventions. GPDS also improves global, regional, and country diarrheal disease burden estimates, informs new enteric vaccine development, and potentially provides a platform for future enteric vaccine evaluation.

## Methods

### Surveillance network design

GPDS builds upon the established GRSN sentinel-site surveillance and tiered laboratory and surveillance system to identify the causes of hospitalized diarrhea in a representative set of LMICs, using a standardized protocol for qPCR testing for a broad panel of diarrheal etiologies. This concept of using existing surveillance infrastructure to capture data on additional pathogens supports WHO’s global strategy for comprehensive vaccine-preventable disease surveillance (7).

### Sentinel surveillance sites

High-performing sentinel surveillance sites from the broader GRSN were chosen to participate in GPDS. These sites were selected to be broadly geographically representative of LMICs, aiming for sites with a history of uninterrupted surveillance and minimum enrollment of 100 cases per year of hospitalized diarrhea in children aged <5 years. To join GPDS, the identified surveillance sites expanded their case definition from only acute, watery diarrhea (the case definition used for rotavirus surveillance) to prospectively enroll all children admitted with diarrhea, regardless of duration or the presence of blood in the stool (8). This approach was initially piloted in 2013-2014 in 16 countries (8), and subsequently implemented and expanded.

Ultimately, 38 sentinel surveillance sites in 31 countries across all 6 WHO regions participated in GPDS during 2017 through 2022 (Figure 1; Table 1). Some countries had only one surveillance site with all patients enrolled from a single sentinel surveillance hospital. Other countries aggregated patients from two or three surveillance hospitals and reported as a single site. Three large countries had multiple GPDS surveillance sites in geographically diverse areas: China (3 sites), India (5 sites)(9), and Pakistan (2 sites). Additionally, while many GPDS sites have participated throughout the entire period of 2017 to 2022, some sites participated for a subset of this time period (Table 1). The numbers of cases enrolled at each site and tested via qPCR reported in Table 1 were calculated using GPDS surveillance data available as of 16 January 2026.

**Fig 1.**
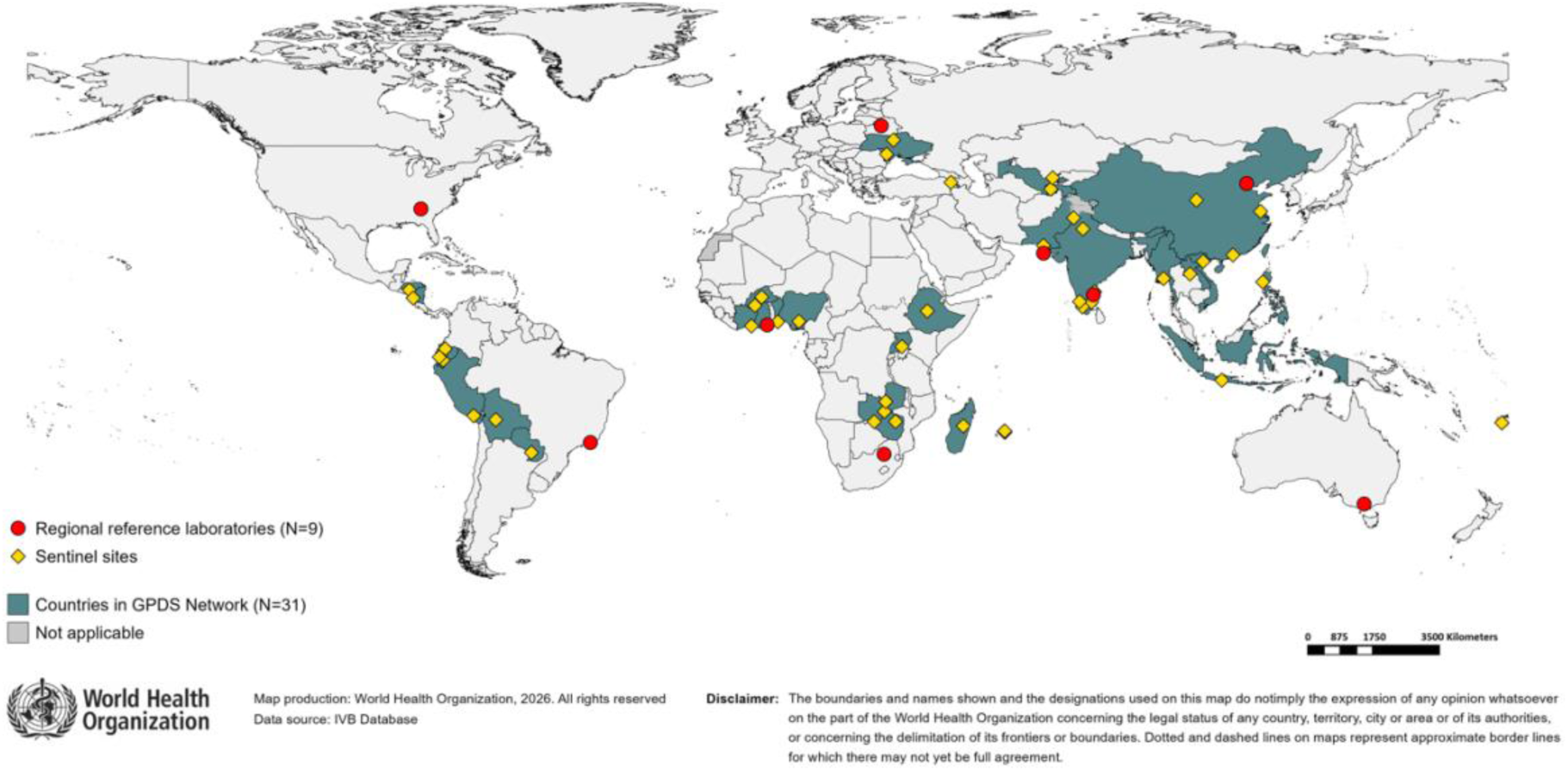
The 9 regional reference laboratories and 38 surveillance sites from 31 countries that participated in Global Pediatric Diarrhea Surveillance during 2017-2022.

**Table 1.**
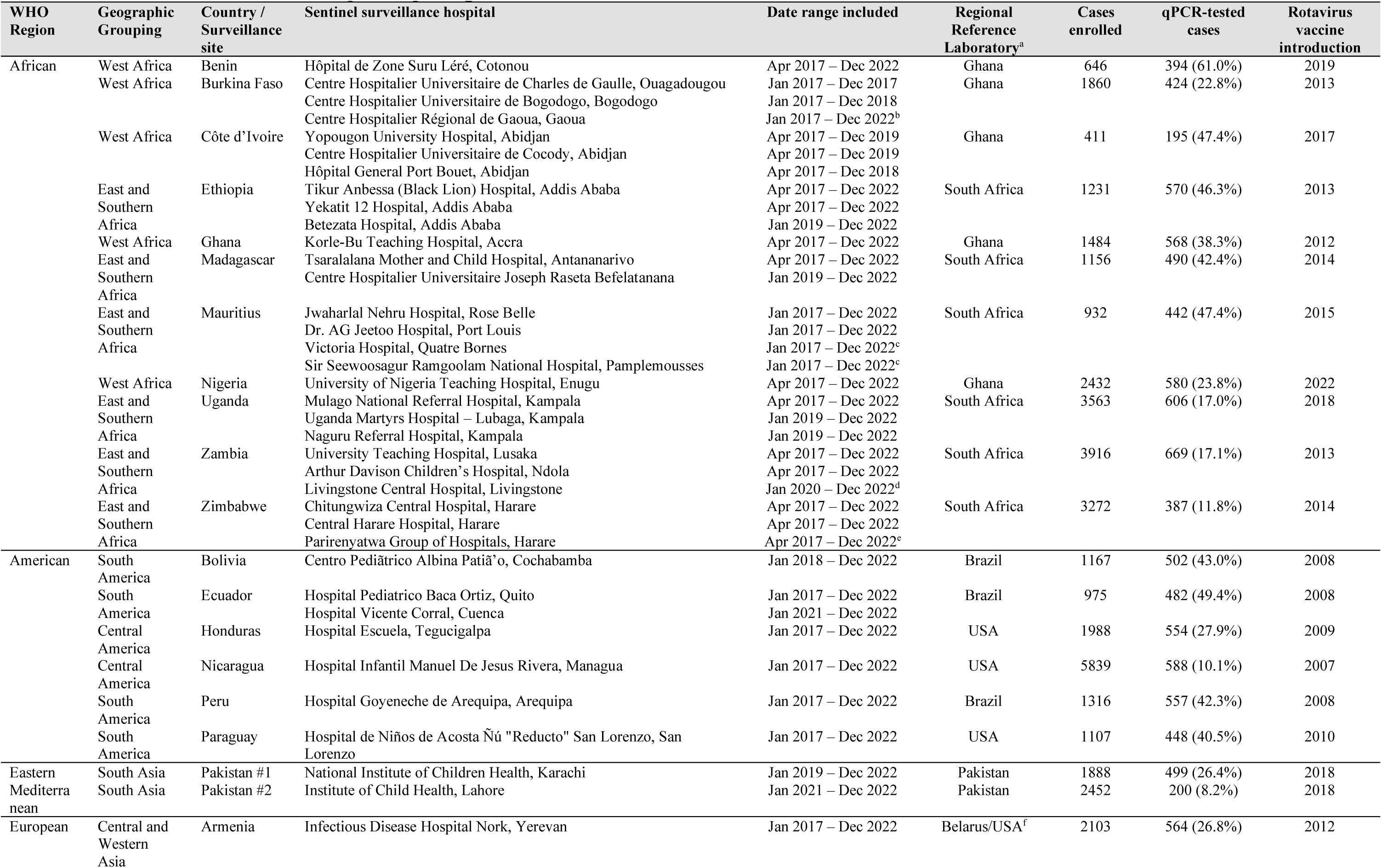
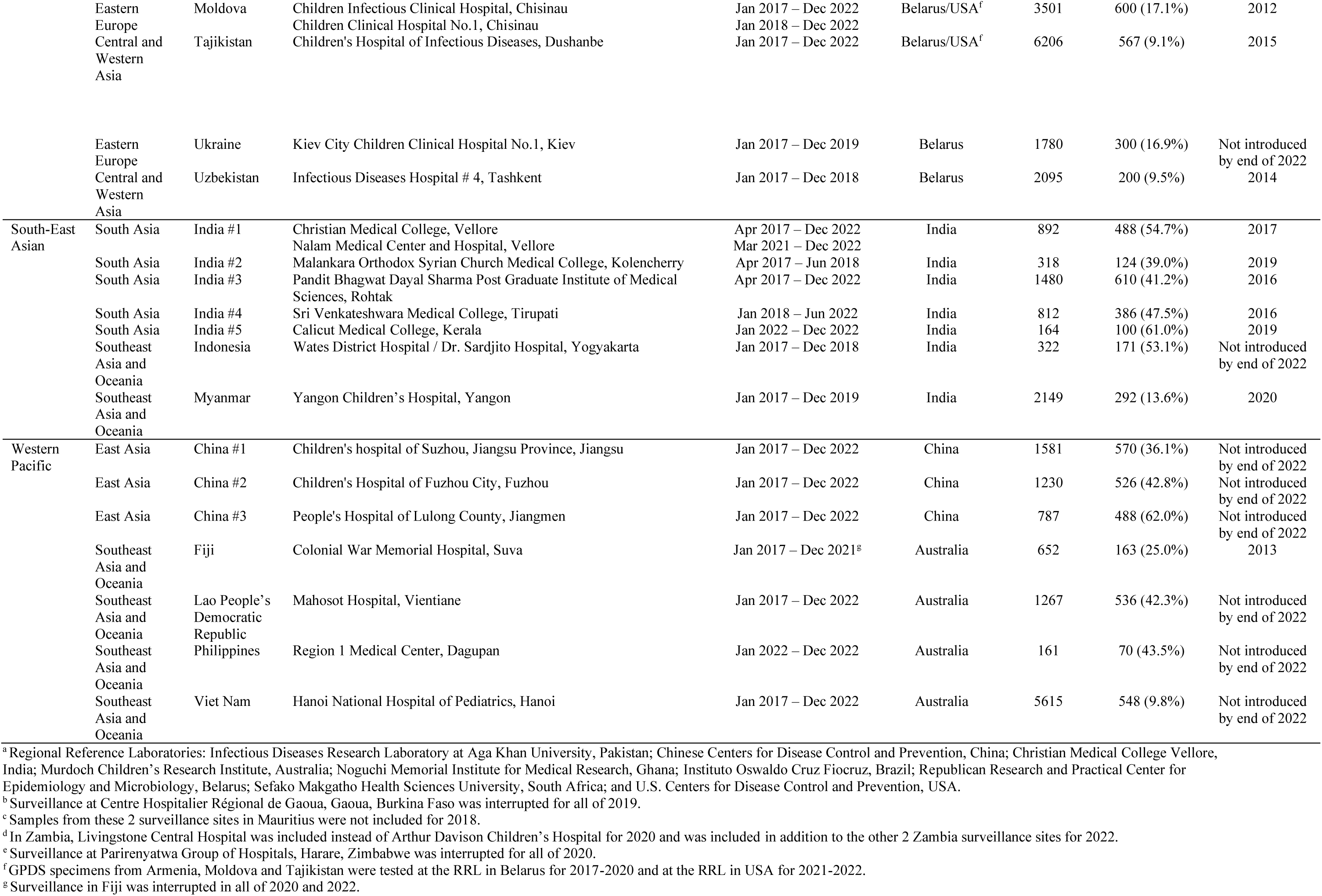
Sentinel surveillance sites participating in Global Pediatric Diarrhea Surveillance, 2017-2022.

### Case definition and enrollment

The inclusion criteria for enrollment in GPDS was any child aged 0-59 months hospitalized with diarrhea at one of the participating sentinel surveillance hospitals. Diarrhea was defined as 3 or more loose stools in a 24-hour period. Children transferred from another hospital or who developed diarrhea during their hospitalization were excluded. Caregivers of children enrolled in GPDS were asked about demographic and clinical characteristics of the cases; medical records were also reviewed. An acute diarrhea episode was defined as an illness with a duration prior to enrollment of fewer than 14 days (through a threshold of <7 days was used in some participating countries), while longer episodes were considered persistent. Additional clinical variables documented reported symptom duration, reported or observed presence of blood in stool, vomiting, dehydration, whether rehydration therapy was given, and whether death occurred in hospital. De-identified data was used to maintain the anonymity of patients.

### Specimen collection, storage, and selection for qPCR testing

Stool specimen collection was attempted from all enrolled cases. Specimens were stored at a maximum temperature of −20°C and at −80°C when possible. All specimens were tested for rotavirus using a commercial enzyme-linked immunosorbent assay (ELISA) as previously described (5). For 2017 and 2018, 25 cases from each surveillance site were randomly selected for qPCR testing for each three-month quarter, as previously described (6). From 2019 through 2022, 100 cases from each surveillance site were randomly selected for qPCR testing at the end of each surveillance year, to simplify sample shipment logistics. Whole stool aliquots were then shipped on wet or dry ice packs to a RRL and stored at −80°C prior to testing.

### Laboratory methods

The RRLs that participated in GPDS testing were Infectious Diseases Research Laboratory at Aga Khan University, Pakistan; Chinese Centers for Disease Control and Prevention, China; Christian Medical College Vellore, India; Murdoch Children’s Research Institute, Australia; Noguchi Memorial Institute for Medical Research, Ghana; Instituto Oswaldo Cruz Fiocruz, Brazil; Republican Research and Practical Center for Epidemiology and Microbiology, Belarus; Sefako Makgatho Health Sciences University, South Africa; and U.S. Centers for Disease Control and Prevention, USA (Table 1). All RRLs participated in site assessments and on-site training as well as annual external quality assurance and quality control exercises (10).

Total nucleic acid extraction was performed as previously described with the Qiagen QIAamp Fast DNA Stool Mini kit (Qiagen, Hilden, Germany) using a modified protocol involving bead beating, with nucleic acid samples subsequently stored at −80°C prior to testing (8). All samples were spiked with Phocine herpes virus (PhHV) and MS2 phage to be used as external controls for DNA and RNA, respectively, and an extraction blank was included in each extraction batch to monitor for contamination. Sample testing using custom-designed TAC qPCR cards (Thermo Fisher, Waltham, MA, USA) was performed as previously described (8). The array card included qPCR assays for the 16 enteric pathogens that were associated with pediatric diarrhea in two prior multisite studies that incorporated diarrheal cases and non-diarrheal controls (Table 2): the Global Enteric Multicenter Study (GEMS) and the Etiology, Risk Factors, and Interactions of Enteric Infections and Malnutrition and the Consequences for Child Health and Development (MAL-ED) birth cohort study (11,12).

**Table 2.**
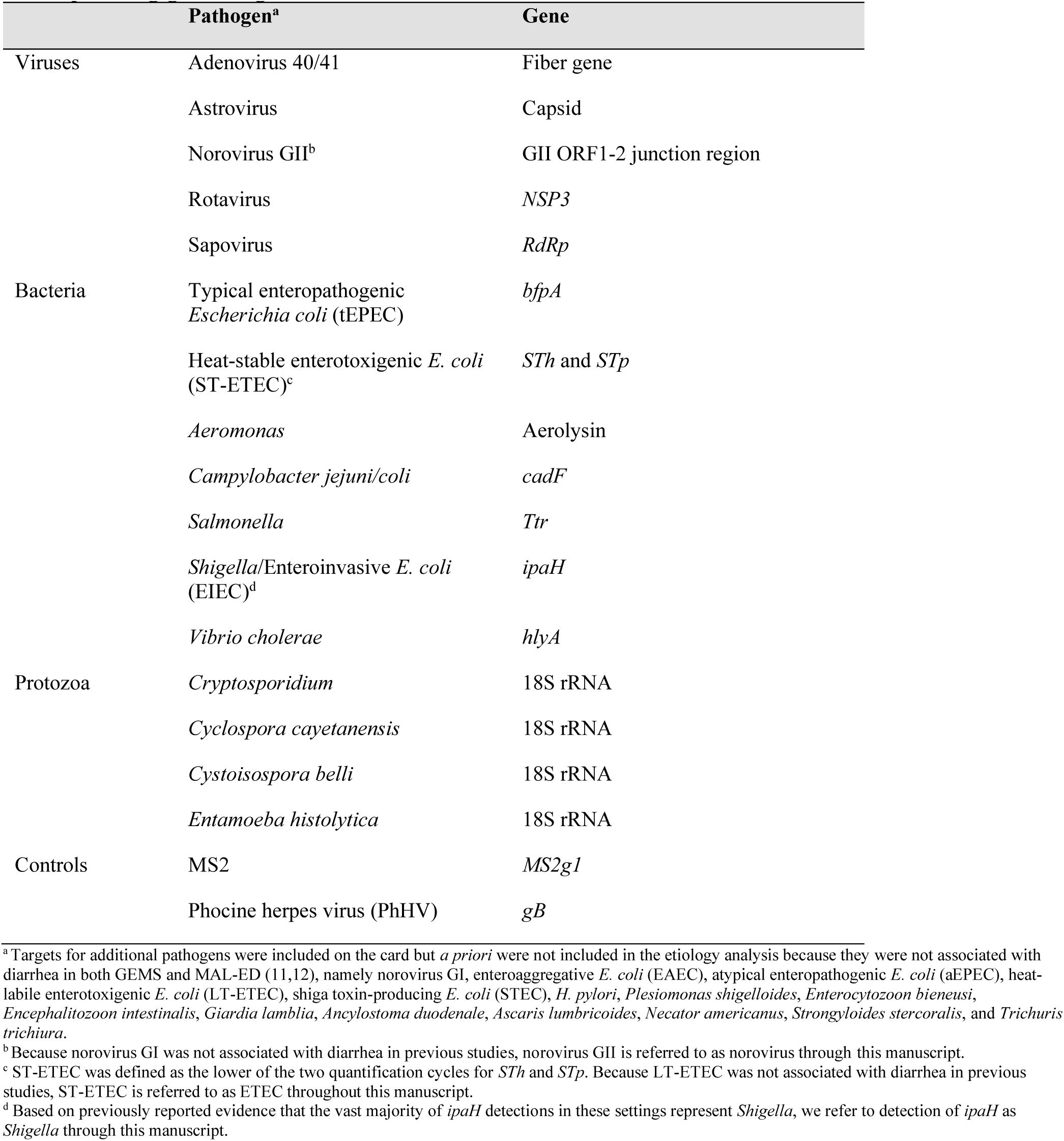
Testing for established etiologies of diarrhea by qPCR on the TaqMan Array Card and corresponding gene target, Global Pediatric Diarrhea Surveillance, 2017-2022.

Raw data analysis was carried out at each of the RRLs with fluorescence thresholds standardized for each qPCR assay between all nine RRLs using software appropriate to each laboratory’s instrument. Beginning with 2020 data, to simplify the process and improve reproducibility, we automated the post-run analysis using machine-learning models, which yielded highly concordant results compared to manual analysis (13). Detections with a cycle threshold (Ct) value less than 35 were considered positive. Negative results were valid only when the corresponding external control was positive, while positive results were valid only when the corresponding extraction blank was negative for the target.

### Statistical analysis

Because samples were selected for qPCR testing by random selection, we applied inverse probability of selection weights such that cases selected for testing were representative of all enrolled cases and appropriately captured seasonal variation in diarrhea etiology. Specifically, the probability of selection for TAC testing was estimated using a logistic regression model which included three-way interactions between study site, dataset, and the quarter and year of sampling as well as between site, dataset, and rotavirus ELISA result. Age in months using linear and quadratic terms and presence of vomiting were also included in the model. To address missing covariate data (specifically ELISA results), we performed Multiple Imputation by Chained Equations (MICE), generating 10 imputed datasets. All covariates included in the weighting model were included in the imputation, in addition to the rotavirus PCR Ct value for the subset of samples tested by TAC. For each dataset, inverse probability weights were calculated from the propensity scores, trimmed at the 99th percentile, and stabilized by normalizing to the mean. The final weights for analysis were derived by averaging the stabilized weights across the 10 imputation iterations for each participant, with untested individuals assigned a weight of zero. These weights were applied to all study estimates that used the subset of tested samples, including clinical characteristics, pathogen prevalence, attributable fractions (AFs), and incidence of hospitalized diarrhea. Analytic results were calculated for two-year periods: 2017-2018, 2019-2020, and 2021-2022. To calculate overall AF estimates for the entire 2017–2022 period, we computed a weighted average of the three period-specific AF point estimates based on the total hospitalized diarrhea episodes in all participating countries in each period. We derived the corresponding 95% confidence intervals by pooling the period-specific variances.

Detection of enteropathogens in stool samples from children without diarrhea symptoms is common in these settings, especially when using molecular testing methods, and non-diarrheal controls were not collected as part of GPDS. Therefore, we attributed diarrhea to specific enteropathogens based on pathogen quantity using models developed from qPCR re-analyses of two multisite etiologic studies of diarrhea that incorporated non-diarrheal controls, the GEMS case-control study and the MAL-ED birth cohort study (11,12,14). Model development for those studies has previously been described in detail (11,12). Briefly, using a conditional logistic regression model for GEMS and a generalized linear mixed-effects model with a random effect for each individual for MAL-ED, a model was fit for each pathogen to describe the association between pathogen quantity and diarrhea, with a random slope for site to allow for variation in the strength of association between pathogen quantity and diarrhea. The MAL-ED model was additionally adjusted for sex, age, and TAC card batch. Pathogen quantity was defined as the log10 increase in pathogen quantity above the analytical cutoff based on the Ct, namely 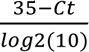 A weighted population AF for each pathogen was then calculated for any stratum of *j* cases as AF = 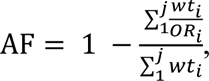 where *wt* is the episode-specific inverse probability weight, and *OR* is the episode-and quantity-specific odds ratio derived from the regression model (15). We propagated uncertainty from the AF estimates using a Monte Carlo approach which simulated 1000 new odds ratio estimates from a normal distribution with the mean derived from the model coefficients and variance-covariance from the covariance matrix (1). To optimize the alignment between GPDS sites and the sites in which attribution modeling had been previously performed, these 1000 estimates were obtained using site-specific model coefficients from GEMS and MAL-ED with a draw distribution determined by weights that minimized the root mean square error distance between the pathogen density distribution in cases at each GPDS site and the proportionally weighted aggregate distribution across the GEMS and MAL-ED sites. In a separate sensitivity analysis, 1000 AF estimates were obtained evenly from the 15 site-specific model coefficients.

To estimate diarrhea incidence, we used national-level estimates of both the incidence of diarrhea hospitalizations and the population of children aged <5 years from the Global Burden of Disease (GBD) 2021 study (1); we used the GBD 2021 estimates for the first year of each 2-year analytic grouping: 2017, 2019, and 2021. To estimate diarrhea hospitalizations, we used the incidence of hospital admissions from International Classification of Diseases (ICD) codes due to diarrhea from 67 sources and 42 countries and found the ratio of hospital admissions to all diarrhea episodes. This proportion represented the proportion of diarrhea for which hospitalization was sought. We modelled this value using DisMod-MR 2.1, a hierarchical Bayesian meta-regression model with the healthcare access and quality index as a predictor (1,16). We then estimated the proportion of diarrhea episodes in children aged <5 years that were hospitalized for each GPDS country and multiplied these by diarrhea incidence to obtain the incidence of hospitalized diarrhea. To estimate variance for all of these estimates, 1000 draw values were obtained from the posterior distribution and propagated through the analyses.

To obtain AF or attributable incidence (AI) estimates across multiple countries, country-level estimates were aggregated. First, for countries with more than one surveillance site, AFs were combined as the mean of each of the 1000 estimates, save for estimates stratified by vaccination status, where AF estimates from India were included independently from each site due to state-level rotavirus vaccine introduction. Then, for each country and pathogen, we calculated the number of pathogen-attributable hospitalizations as the product of one randomly-sampled draw of the country-level AF and one randomly-sampled draw of the number of diarrhea hospitalizations; this was repeated 10,000 times. Each estimate was summed across all included countries to obtain the total number of attributable episodes for each pathogen and was then either divided by the under 5 population denominator to obtain AI estimates or divided by the total number of diarrhea hospitalizations to obtain AF estimates. An analogous analysis was conducted to obtain attributable episodes.

We considered rotavirus vaccines to be introduced at each site if rotavirus vaccination had been added to the immunization program at the country or subnational level by the end of first year of each 2-year analytic grouping (3). In addition to the 6 WHO Regions, analytic geographic groupings were created to provide greater distinction and public health inferences for the GPDS estimates; see Table 1 for the geographic grouping assigned to each country and surveillance site.

For cases where multiple pathogens were detected in the stool sample, we determined the single most likely etiologic pathogen by calculating the individual etiologic attributable fraction (AF_e_) for each pathogen and then choosing the pathogen with the highest AF_e_ (as long as the AF_e_ was >0.2). The AF_e_ represents the individual attributable fraction for each pathogen within each diarrheal episode, serving as an episode- and pathogen quantity-specific metric of attribution of that pathogen as the cause of the illness, with possible values ranging from 0 to 1, where 1 is “fully attributable”. These values were generated by calculating the mean of 100 individual AF_e_ estimates produced from a weighted draw distribution (described above) of the GEMS and MAL-ED attribution models. The AF_e_ for a specific pathogen in an episode is calculated as 1 - 1/*OR*, where *OR* is the pathogen- and quantity-specific odds ratio.

A diarrhea severity score (see Table 3) with a maximum possible score of 16 was derived for each enrolled case using the Modified Vesikari Score (MVS) (17) as a starting point but with two important modifications. First, we omitted child temperature because these data were not available. Second, because GPDS explicitly enrolls hospitalized children, we compressed the points for dehydration, such that some and severe dehydration received 1 and 2 points respectively, rather than 2 and 3 points per the MVS. We applied the standard MVS severity cutoffs to the GPDS severity score. Although the maximum score was reduced, the restriction of our cohort to hospitalized children meant the severity distribution remained skewed high (16.9% Mild, 41.4% Moderate, and 41.7% Severe), supporting the use of the original cutoffs.

**Table 3.** Diarrhea severity score derived for the Global Pediatric Diarrhea Surveillance network.

| Score component | Assigned points |
| --- | --- |
| Duration of diarrhea | 1–4 days (1 pt)<br>5 days (2 pts)<br>≥ 6 days (3 pts) |
| Max stool frequency (per 24h) | 1–3 stools (1 pt)<br>4–5 stools (2 pts)<br>≥ 6 stools (3 pts) |
| Duration of vomiting | 1 day (1 pt)<br>2 days (2 pts)<br>≥ 3 days (3 pts) |
| Max vomiting episodes (per 24h) | 1 episode (1 pt)<br>2–4 episodes (2 pts)<br>≥ 5 episodes (3 pts) |
| Dehydration | None (0 pts)<br>Some (1 pt)<br>Severe (2 pts) |
| Treatment | None (0 pts)<br>Oral Rehydration (1 pt)<br>IV Rehydration (2 pts) |
| Score categories | Mild illness (0–6 points)<br>Moderate illness (7–10 points)<br>Severe illness (11–16 points)<br>[Total out of 16 points] |

To describe the association between pathogen attribution and clinical characteristics of each diarrhea episode, we modeled the association between the pathogen AF_e_ and the clinical characteristic using a Poisson regression model for binary outcomes and a linear regression model for continuous outcomes. We calculated risk ratios for associations with binary variables and mean differences for continuous variables. All models were adjusted for the site’s geographic grouping (see Table 1) and the year of enrollment.

To evaluate for seasonal trends in pathogen-specific diarrhea, for each geographic grouping, we used Poisson harmonic regressions to estimate pathogen-attributable epsiodes as a function of the calendar week (*w*) of enrollment using the terms 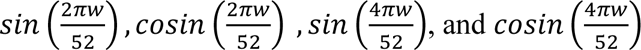. We then used this model to predict the weekly incidence of pathogen-specific diarrhea.

### Ethical considerations

Surveillance activities that are part of GPDS are exempt from research ethical review by WHO, since they are considered public health surveillance.

## Strengths and limitations

GPDS makes a major step forward in filling the data gap in our understanding of the full range of severe childhood diarrheal etiologies at a global, regional, and country level. By incorporating quantitative molecular testing and pathogen attribution modelling into a global network with standardized surveillance, sample collection and testing, and analytic protocols, GPDS provides direct estimates of pathogens causing hospitalized diarrhea in children aged <5 years in LMICs in 2017–2022. By design, GPDS includes a large, globally diverse selection of LMICs, including countries and geographies that are not well represented in the published literature. As the 31 LMICs included in GPDS represented more than 60% of incident diarrhea and diarrheal deaths in 2021, the aggregated results are broadly representative of all LMICs. GPDS data feeds back into global disease burden estimates, helping to improve these estimates over time (1,18).

Despite these clear strengths of GPDS, the resulting analytic estimates do have some limitations. To begin, GPDS focuses on the most severe subset of diarrhea, by enrolling hospitalized children. Though estimates were adjusted for the proportion of diarrheal cases requiring hospitalization in various settings, the results may not be fully representative of the etiologies causing milder diarrhea. Next, GPDS sentinel surveillance hospitals are predominantly but not exclusively referral hospitals in urban centers and thus may not be representative of all hospitalized diarrhea across each country. Similarly, some GPDS regional estimates come from one or a few countries and thus may not be representative of all countries in the area. Also, sentinel surveillance networks such as GPDS are not designed for outbreak detection or to fully represent the burden of diarrhea due to outbreak-prone pathogens, such as cholera. Additionally, GPDS does not enroll concurrent controls, but rather uses attribution models developed in two previous multisite pediatric diarrhea etiology studies, GEMS and MAL-ED, which were performed across 15 sites in Asia, Africa, and South America from 2007-2012 using the same diagnostic platform, but not part of GPDS (11,12,14). As GPDS sites do not have defined catchment areas, our extrapolation to incidence of diarrhea hospitalizations and deaths relied on national GBD model estimates. Finally, individual vaccination status was not available for all enrolled patients, nor was vaccination coverage in individual site catchment areas, thus we could not directly evaluate vaccine impact but can rather look at the broad impacts of vaccine introduction at a population level over time.

## Conclusions

By leveraging existing rotavirus sentinel surveillance in a globally representative range of LMICs and applying the best-available diagnostic and analytic methods, GPDS provides a broad and detailed picture of the etiology of hospitalized diarrhea in children aged <5 years in the post-rotavirus vaccine era. The network monitors the changing burden of pathogens over time, monitors circulating strains, and generates data to inform decision-making around public health interventions. GPDS also improves diarrheal disease burden estimates at the country, regional, and global levels and informs new enteric vaccine development, such as for *Shigella* and norovirus.

## Data Availability

Aggregate GPDS data is available online at: https://immunizationdata.who.int/global?topic=Rotavirus-and-pediatric-diarrhea-surveillance-data. De-identified participant-level data used in these analyses will be made available by WHO upon request to qualified researchers, after approval of a proposal submitted to and signing of a WHO data sharing agreement.

## Acknowledgements

We would like to thank the sentinel surveillance hospitals and staff, national laboratories, and the country Ministries of Health for supporting and maintaining surveillance at the country level, as well as the WHO Country Offices. The findings and conclusions of this report are those of the authors and do not necessarily represent the official position of the U.S. Centers for Disease Control and Prevention or the World Health Organization.

## Appendix. Global Pediatric Diarrhea Surveillance network Group Authors

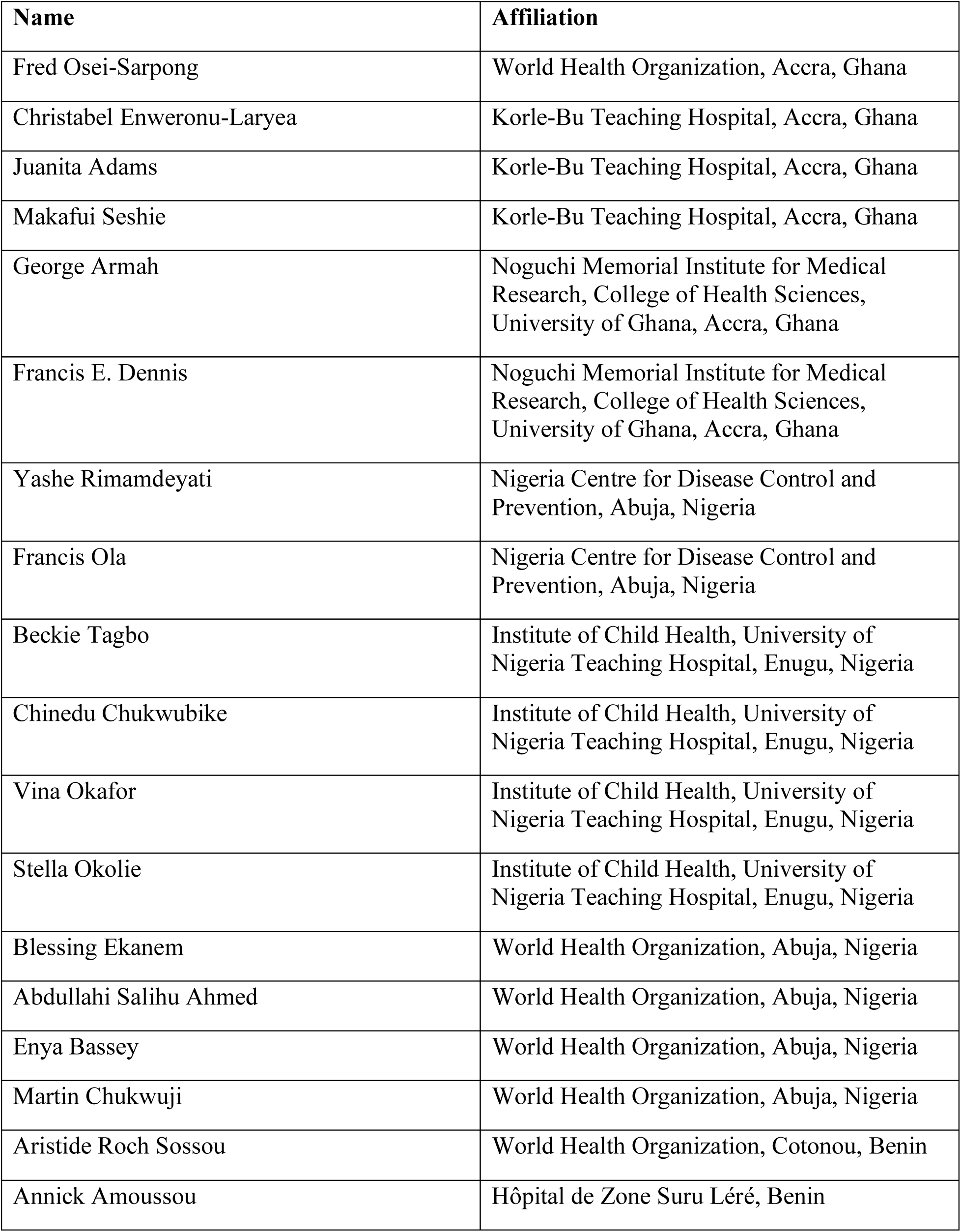
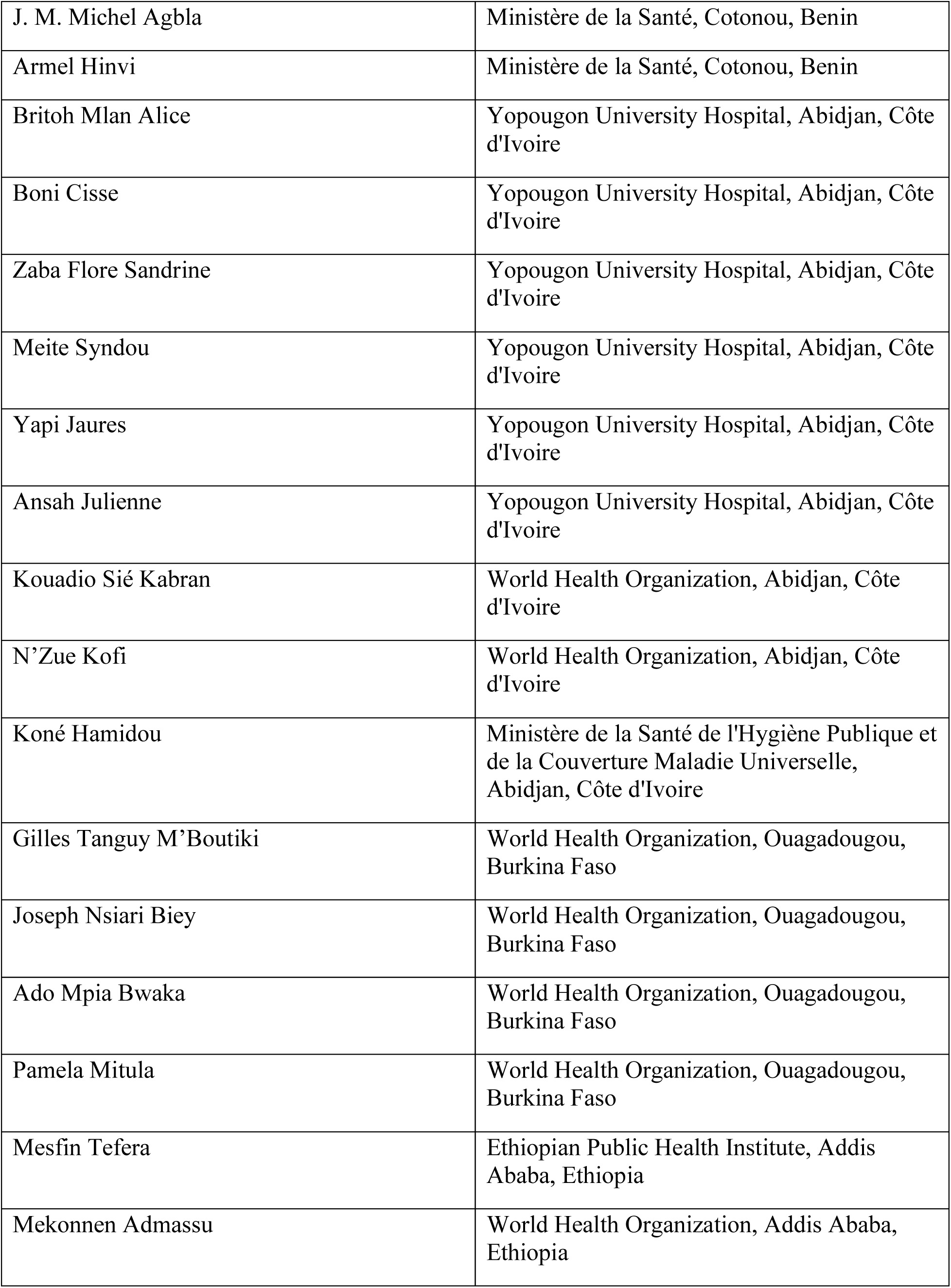
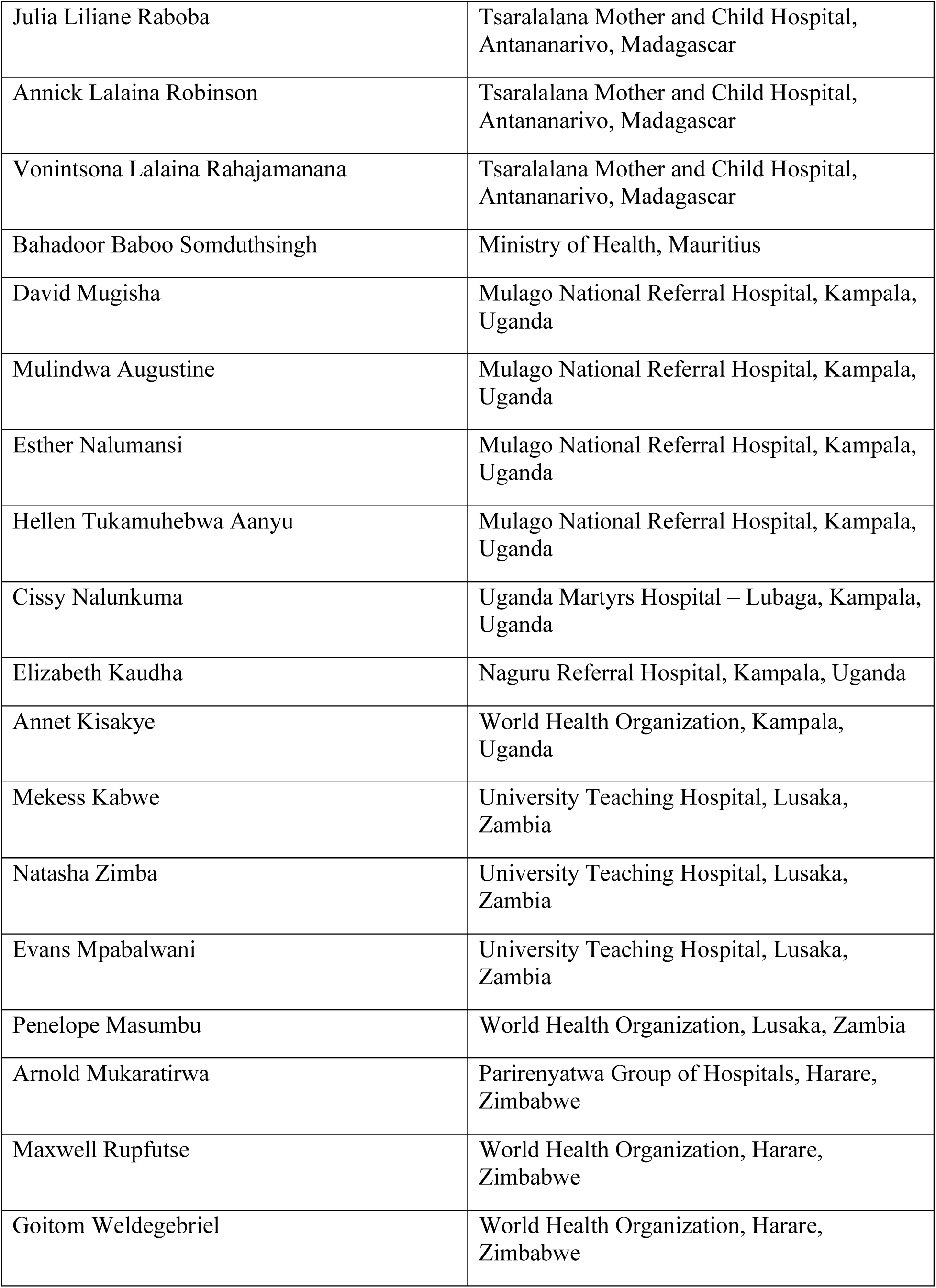
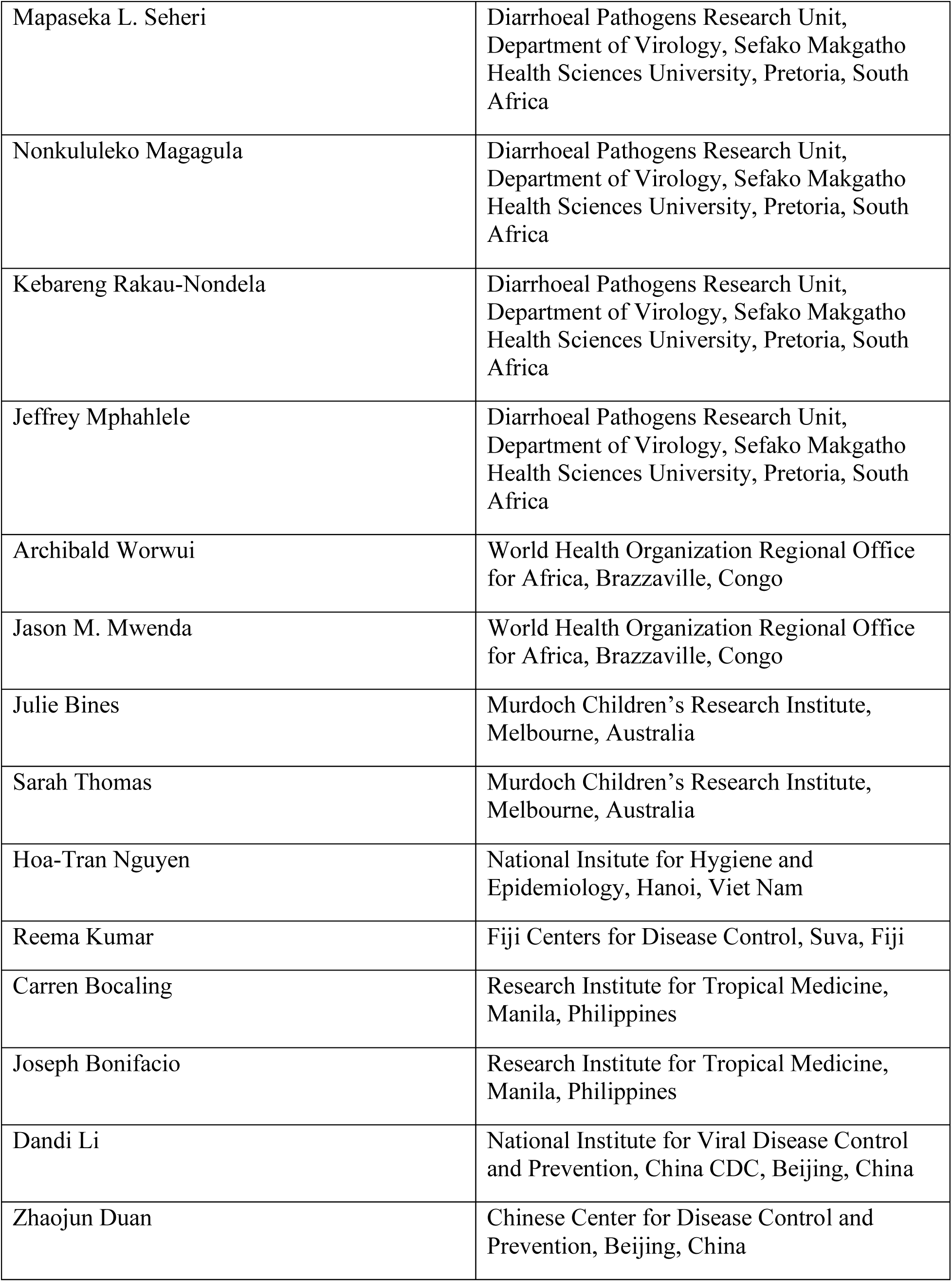
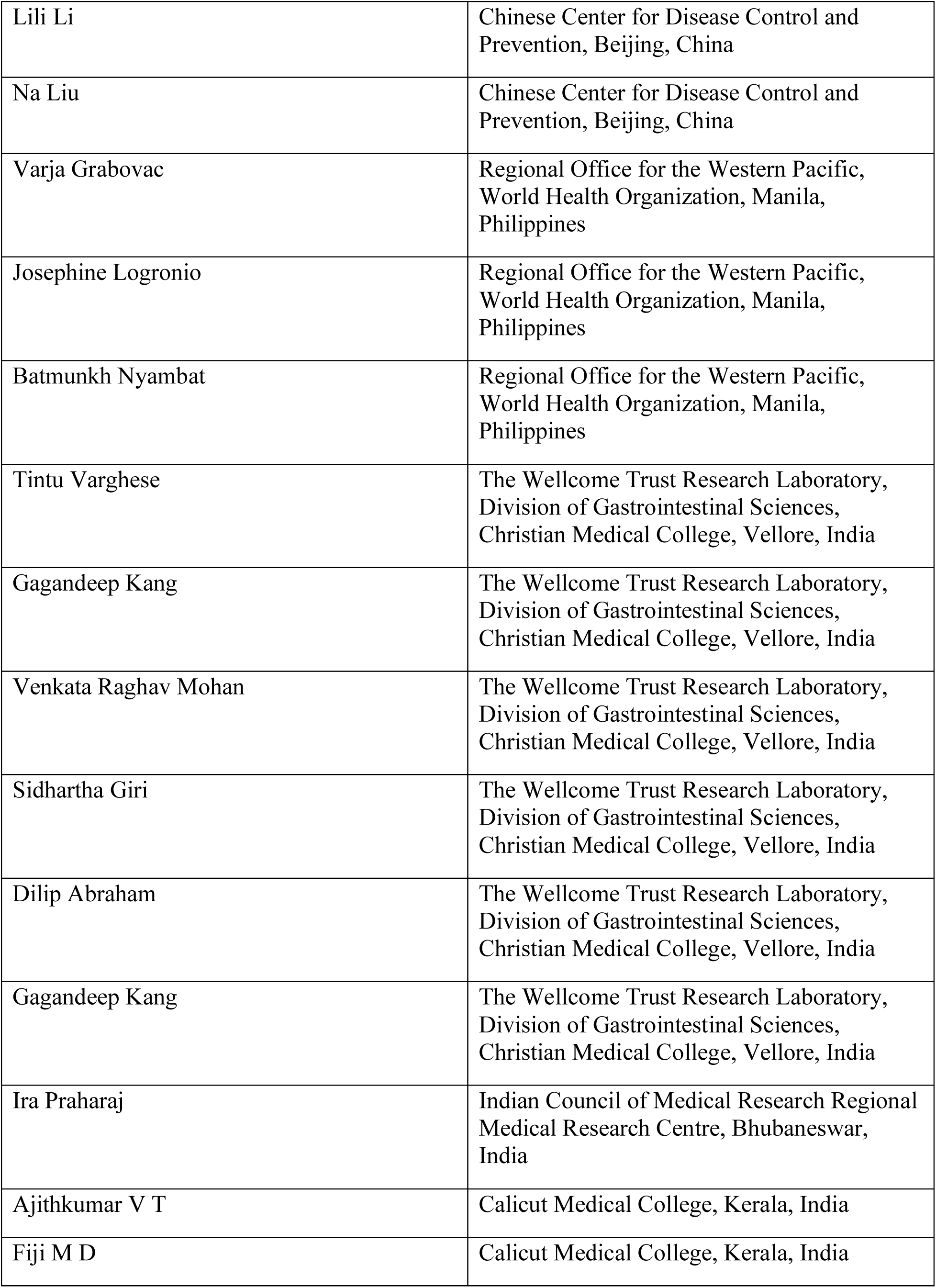
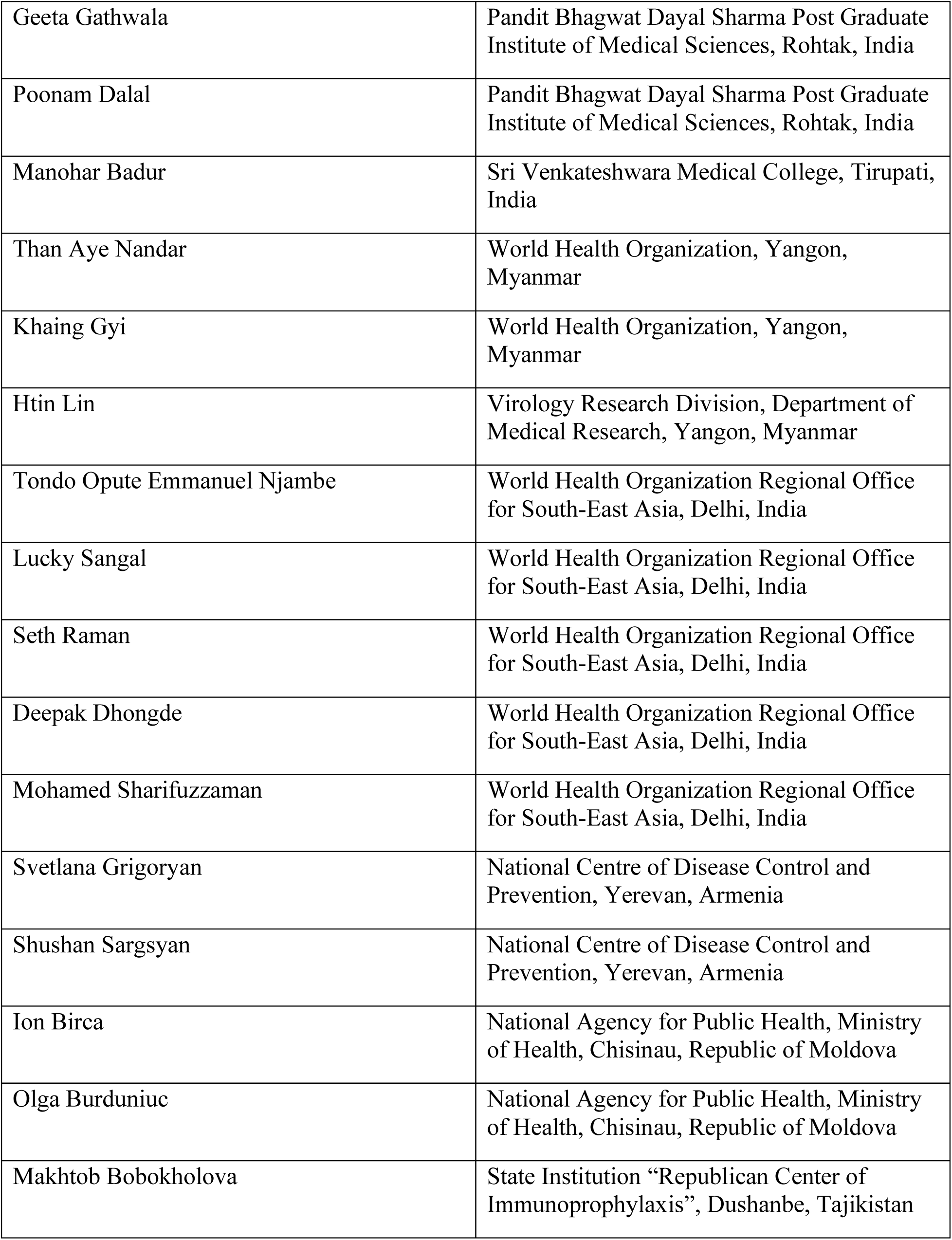
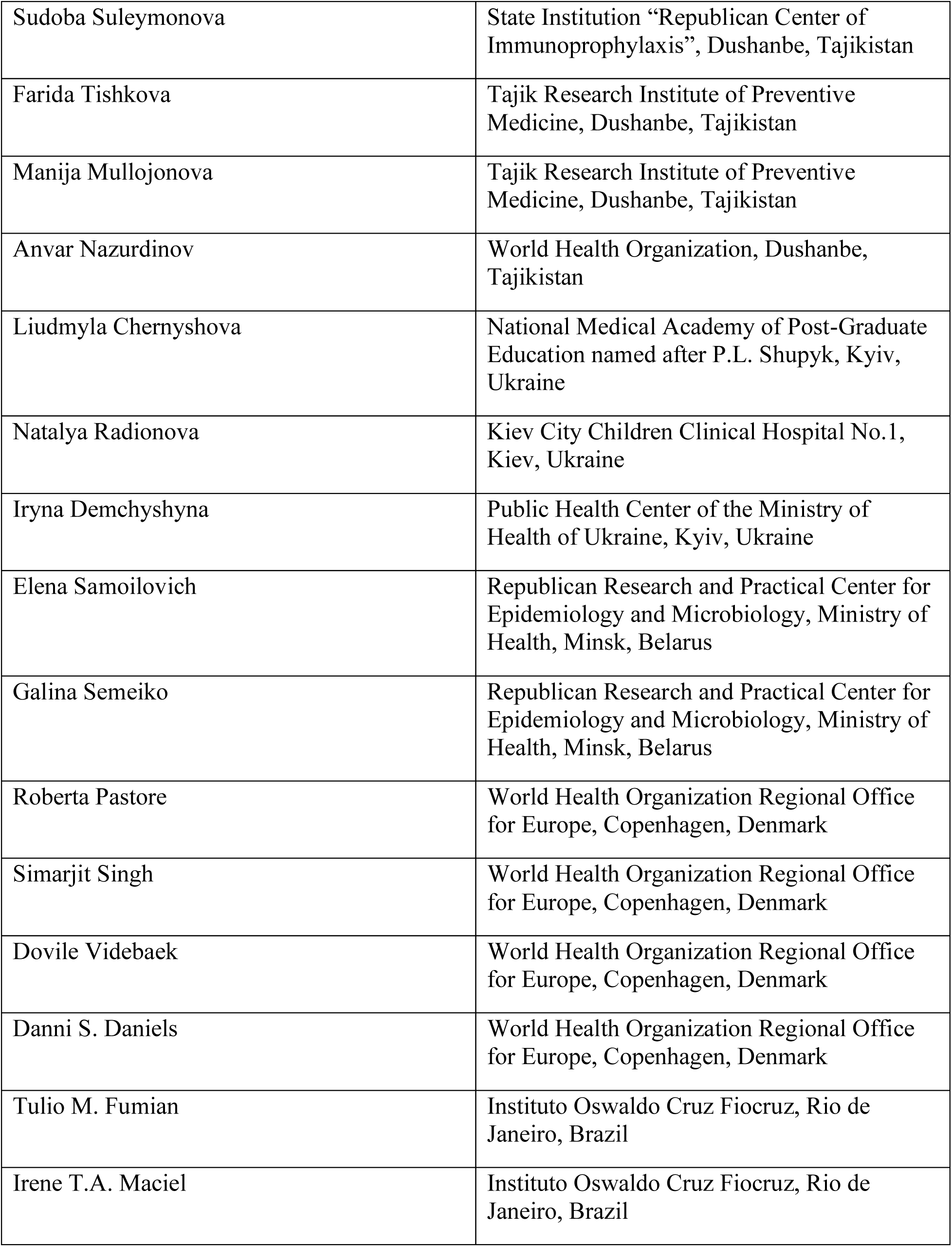
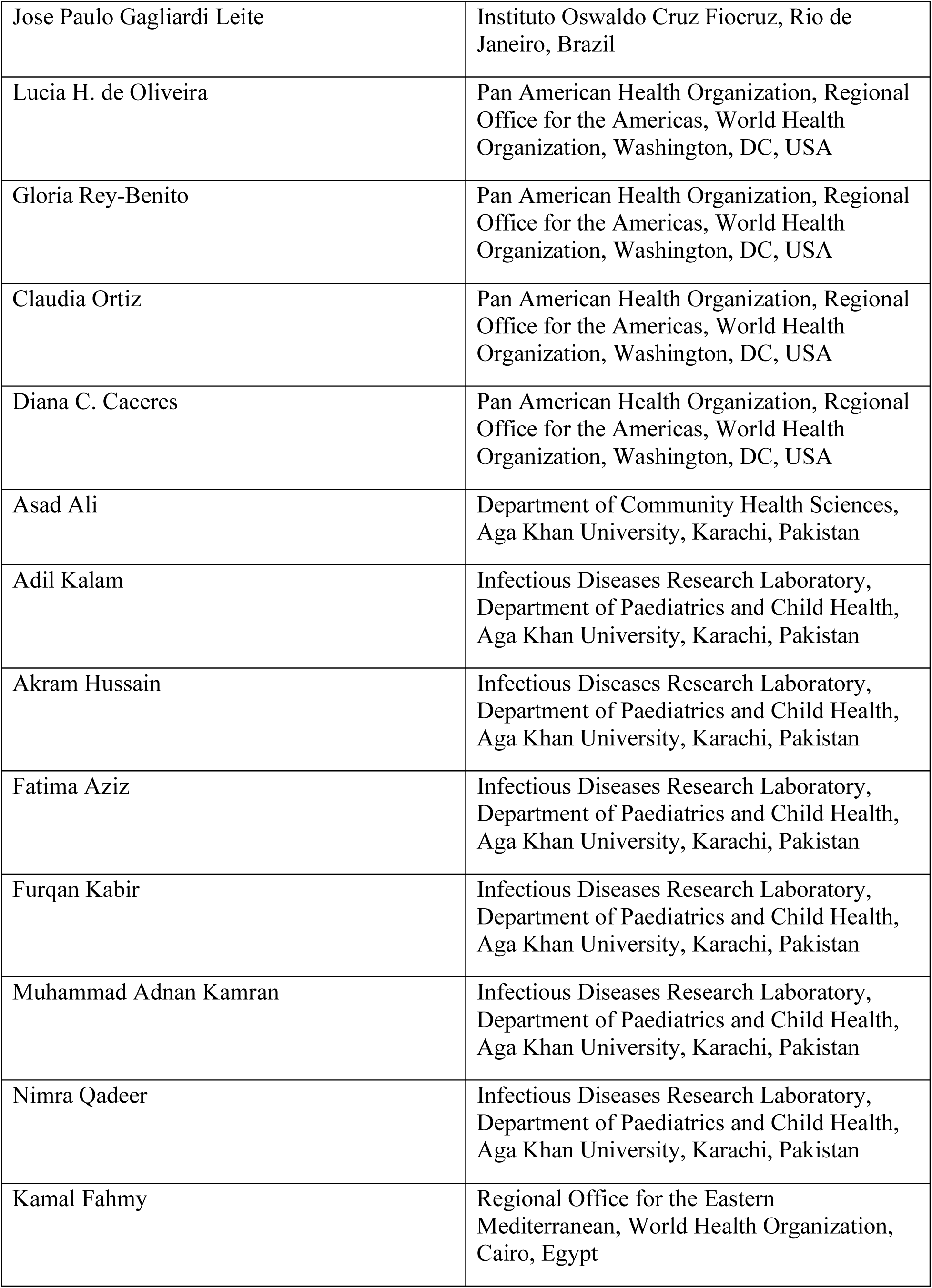
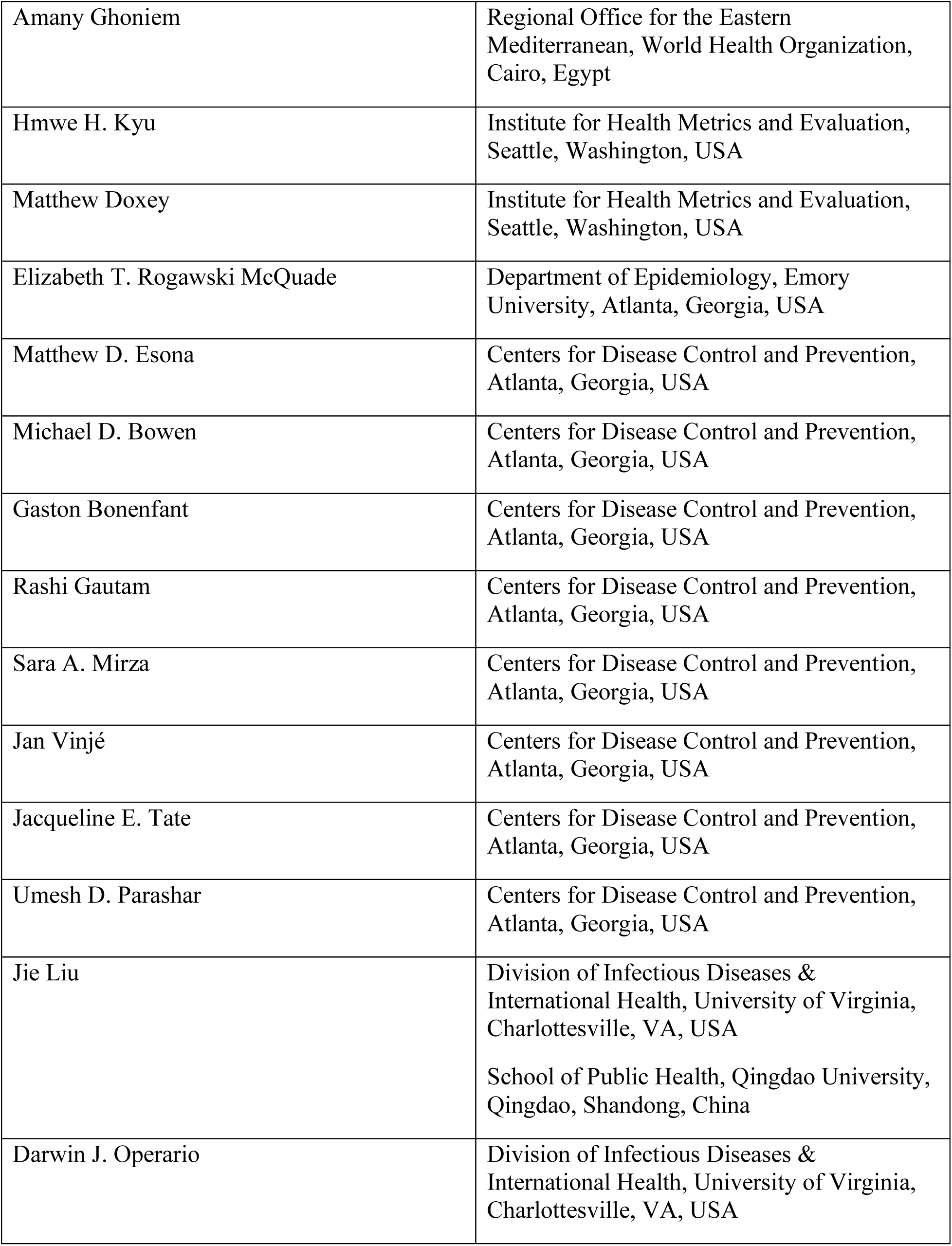
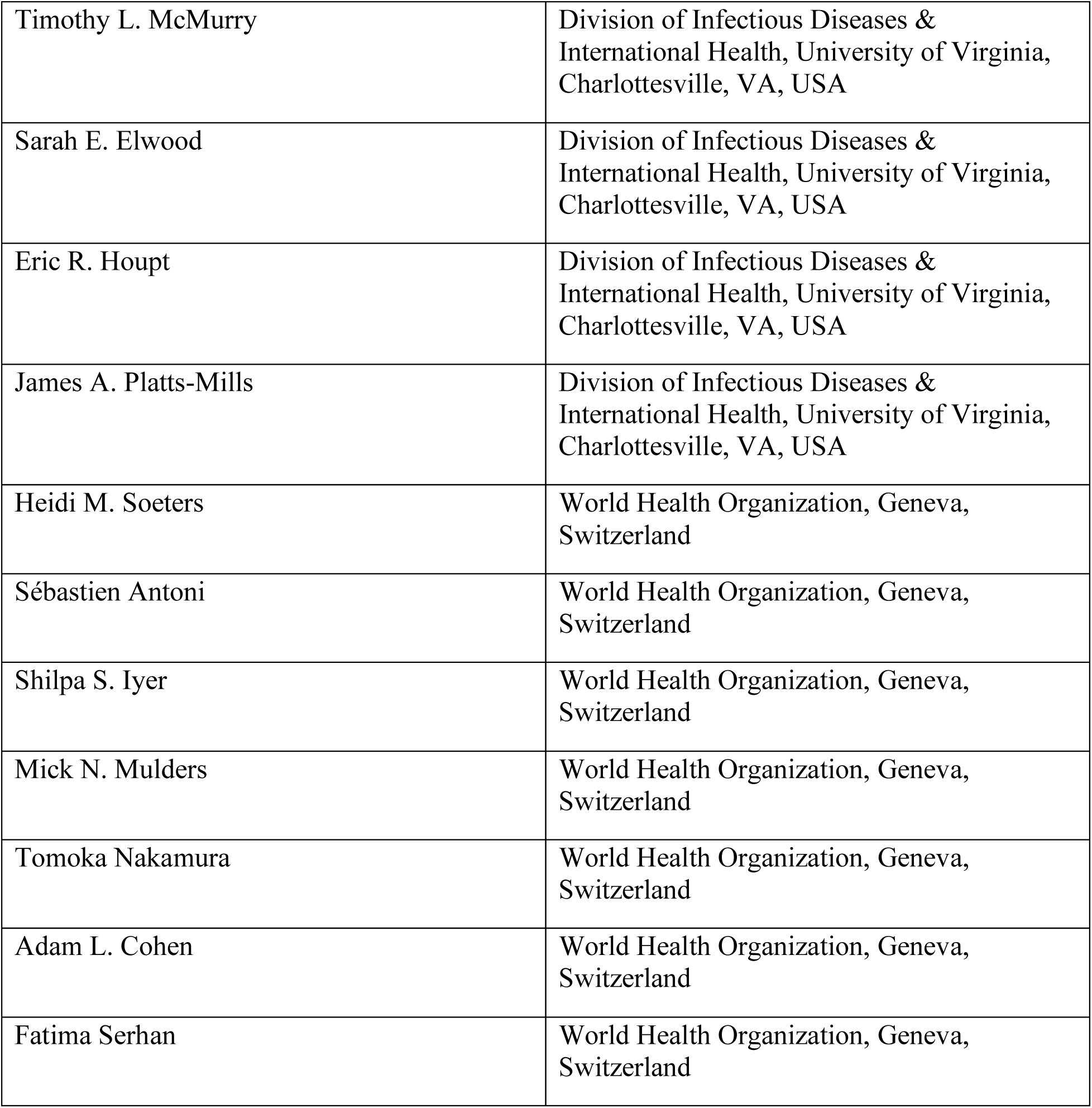

